# Role of Oxygenation Devices in Alleviating the Oxygen Crisis in India

**DOI:** 10.1101/2022.10.15.22281117

**Authors:** Deepshikha Batheja, Vinith Kurian, Sharon Buteau, Neetha Joy, Ajay Nair

**Affiliations:** Post-Doctoral Fellow, One Health Trust (formerly CDDEP), Bengaluru, Karnataka, India; Research Manager, LEAD at Krea University, Chennai, Tamil Nadu, India; Executive Director, LEAD at Krea University, Chennai, Tamil Nadu, India; Program Director, ACT Grants, Bengaluru, Karnataka, India; Chief Executive Officer, Swasth Alliance, Bengaluru, Karnataka, India

**Keywords:** COVID-19, Oxygen concentrators, Oxygen therapy, SARS-CoV-2, India

## Abstract

**Introduction:** There has been an unprecedented increase in global demand for medical oxygen equipment to solve the acute oxygen shortages caused by SARS-CoV-2 infection. The study aims to assess the value of improved access and use of OCs and cylinders during the COVID-19 pandemic in India. This evaluation is relevant to strengthening health systems in many resource-constrained LMIC settings.

**Methods:** Using a Probability Proportional to Size (PPS) sampling method, primary surveys were conducted in 450 health facilities across 21 states in India. The primary outcomes measured were self-reported utility of oxygenation devices in meeting the oxygen demand in the short-run and long-run utility of devices compared to the pre-oxygen-devices-distribution-period. We perform bivariate and multivariate regression analyses.

**Results:** Around 53-54% of surveyed facilities reported that the distributed oxygenation devices helped meet oxygen demand in the short run and are expected to increase their long-run capacity to admit non-COVID patients with oxygen needs. Timely availability of technicians was associated with meeting oxygen demand using the additional oxygenation devices at the facilities. Facilities that increased the number of staff members who were able to administer oxygen devices were at higher odds of reducing the administrative load on their staff to organize oxygen support in the long run. Hospital infrastructure was also associated with the long-run outcomes.

**Conclusion:** We find that oxygenation devices such as cylinders and OCs were useful in addressing the oxygen demand during the COVID-19-related oxygen emergency. Overall production of oxygen to meet the demands and investments in training personnel to administer oxygen could help save lives.

**Funding:** None

**Research in Context:** *What is already known on this topic?:* Oxygen therapy is an essential medicine for the treatment of severely ill patients with COVID-19. Availability of adequate oxygen support was therefore crucial for every health facility that serves COVID-19 patients, particularly in low-resource settings. Medical equipment donation to low-resource settings is also a frequently used strategy to address existing disparities, but there is a paucity of reported experience and evaluation of the impact of these devices. Challenges such as infrastructure gaps, lack of technological and maintenance capabilities, and non-prioritization of essential supplies have previously been highlighted in other developing-country contexts.

*What this study adds?:* Timely availability of technicians, the average load of COVID-19 patients during the second wave, and timely availability of oxygenation devices such as OCs were factors associated with the additional oxygenation devices having a significant impact on meeting the oxygen demand at the facility. Further, facilities that increased the number of staff members that were able to administer oxygen devices at the beginning of the second wave were at higher odds of expecting a reduction in the administrative load on their staff to organize oxygen support in the long run.

*How this study might affect research, practice or policy?:*

- This is the first study to demonstrate the utility of oxygen devices such as cylinders and OCs in meeting oxygen demand during the COVID-19 oxygen emergency.
- Prior findings of the literature from other LMICs stress the importance of hospital infrastructure such as power outlets in the effective use of these oxygen devices. Our analysis also finds these barriers to be significant and additionally suggests that the timely availability of oxygen administrators and technicians is crucial in the utilization of these devices.

## Introduction

The COVID-19 crisis amplified a critical vulnerability in the healthcare infrastructure of many low- and middle-income countries (LMICs)—a severe shortage of medical oxygen (Baker, 2020; Dauncey JW, 2019). Critically ill patients need medical-grade oxygen for invasive ventilation and low- and high-flow oxygen therapies (Duke, Graham, & Cherian, 2010). An estimated half a million people in LMICs need 1.1 million cylinders of oxygen per day, with 25 countries in 2021 reporting surges in demand for medical oxygen up to a degree of shortage multiple of 60, the majority of countries with oxygen deficits are concentrated in Africa, Middle East and South Asia (Howie, 2010; Macnamara, 2020). In 2015, the Lancet Commission on Global Surgery reported that approximately one-quarter of hospitals surveyed in resource-limited countries lack sufficient oxygen supply (Meara, 2015).

On 26th April 2021, India saw the highest daily tally of the new SARS-CoV-2 infections ever recorded at the time in the world, 360,960 cases per day, taking the pandemic total to 16 million cases, globally. Among the myriad complications associated with responding to the crisis, demand for oxygen required for critical care patients significantly exceeded its supply. Hospitals across the country faced a major crisis of short-term, mobile sources of oxygenation, imperative to the health of affected patients, disproportionately increasing mortality rates. As a response to the crisis, the country saw an influx of oxygen cylinders, oxygen concentrators (OC) and setting up of Pressure Swing Adsorption (PSA) plants (Thadani, 2021). Import of oxygen therapy apparatus has increased drastically over the past three years. The first lockdown was announced in end-March 2020 and imports of oxygen apparatus stood at 1.74 million units, which is almost three times of what it was in 2017-18. In 2020-21, the number further went up to 2.5 million units^1^.

While medical equipment donation to low-resource settings is also a frequently used strategy to address existing disparities, there is a paucity of supportive evidence of its utility (Stein, Perry, & Banda, 2020). It is crucial to analyse the benefit of these devices and the associated costs, such as ease of use and cost of maintenance/repair before more investments are made in procuring more oxygenation devices. Some associated challenges such as infrastructure gaps, lack of technological and maintenance capabilities, and non-prioritisation of essential supplies have previously been highlighted in other developing-country contexts (Marks, Thomas, & Bakhet, 2019). We contribute to this literature by assessing the utility and barriers in access and use of OCs and cylinder in the COVID-19 pandemic in the Indian setting during a global emergency. We focus on both short-term and long-run outcomes to explore how donated medical equipment such as oxygenation devices can be used and maintained effectively to achieve a longer life. Such an evaluation is also essential to make policy level changes, as we work towards strengthening the health system to tackle and prevent similar crises in the future in resource-constrained LMIC settings.

## Methodology

The study uses primary survey data from 450 health care facilities across 21 states in India where a philanthropic initiative by ACT Grants and Swasth Alliance had distributed over 50000 oxygen equipment during the second wave.

### Sample Selection

The sampling frame consisted of all hospitals that received oxygenation devices (OCs and/or cylinders) as part of the COVID-19 relief initiatives of ACT grants and Swasth Alliance. This includes a total of 2158 facilities. The sample selection process was conducted in two stages. First, to identify a selection of districts that represent varying level of poverty and COVID-19 caseload intensities. Second, a stratified selection of different types of facilities that received oxygen devices was randomly selected to have representation across different types of facilities ranging from COVID Care Centres, Private facilities and Government facilities of different grades.

#### District Sample Selection

To generate a sample of districts, a combination of three secondary datasets was utilised.

1. District Level COVID-19 Caseload data^2^ (To measure COVID Intensity at a district level)
2. District Level Multi-Poverty Index Data^3^ (To measure intensity of poverty among beneficiary districts)
3. Healthcare Facility MIS Data^4^ (To map Facilities where surveys can be conducted)

Probability Proportional to Size (PPS) sampling method was adopted assigning the proportion of oxygen devices distributed within each district as the variable indicative of size to ensure that proportional representation of states and districts be made in line with the distribution of devices within these districts. A sample of 100 districts was drawn which accounted for 60% of the total equipment distributed.

#### Facility Sample Selection

For the sampled 100 district, a stratified random sampling of 6 facilities each was conducted across the following strata of healthcare facilities where oxygen devices were sent. 1. COVID Care Centres, 2. Government Hospitals (Above the Primary Level), 3. Government PHC/CHC Facilities, 4. Private Facilities, 5. Other Facilities (ex. Military/ Railway Hospitals)

A total of 558 endpoint facilities were selected for the primary data collection exercise. An oversampling of facilities was done to account for non-responses/ refusals in participating in the survey exercise among facilities. A total of 450 healthcare facilities were surveyed in-person between October and December 2021.

### Outcomes Measured

The survey instrument was designed using the SurveyCTO platform and administered using a tablet device in-person. The survey instrument captured questions on the perceived impact of supply of oxygen devices by ACT Grants and Swasth Alliance on COVID-19 response. The following outcomes were used to measure the utility of oxygenation devices in alleviating oxygen supply shortages in the short-run and long-run:

1. Effect of additional delivery of OCs and cylinders on oxygen demand in the short-run was explored using a likert-scale question and combining responses such as “Significantly decreased” and “Completely/fully met” oxygen demand to indicate a positive response.
2. Expected increase in capacity of the facility in the long-run was a self-reported question on whether respondents/facilities would be able to admit non-COVID patients with oxygen needs in future (long-run).
3. Expected reduced administrative load on staff to organize oxygen support in the long-run was a self-reported yes/no question on the same.

### Patient and public involvement statement

We piloted the survey tool among a few hospital administrators, whose inputs helped us refine the survey design.

### Data Analysis

Descriptive, bivariate, and multivariate regression analyses were conducted. Bivariate analyses were performed to identify significant associations between the dependent and independent variables. Those independent variables significantly associated with the dependent variables at a p-value <0·05 were entered into the multivariate model. Since our main outcomes are binary, we use a logistic regression model was used to identify their associations with various independent variables (Odd’s ratio for bivariate analysis, adjusted Odd’s ratio for multivariate analysis). We cluster the standard errors at the state-level. Statistical analyses were performed using STATA v.16® software.

Data on the variables of interest were extracted from the datasets stored in software encrypted files, cleaned and recoded into categorical values. Less than 5% of the data was missing. Missing data was imputed with mean response to the question observed in the overall data.

### Definition of variables

#### Independent variables

We test for associations of dependent variables with independent variables such as the characteristics of the healthcare facilities: type of facility (where comparison/reference category is covid care centre, private and other facilities i.e., all non-government and private entities), geographic location being rural (with urban as the reference category, which includes “urban” and “mostly urban” areas), the availability of oxygenation devices (reference category is facilities that did not receive any OCs or cylinders) and the timeliness of receiving devices with respect to the second wave of COVID-19 at the district level. Self-reported average patient load during the 2nd wave (reference category is “0” patient load) was also included to measure the facility’s capacity to cater to COVID-19 patients. Variables measuring changes to healthcare personnel including the increase in medical equipment administrators and technicians during COVID-19 (reference category is facilities that did not increase number of staff members who can administer oxygen). We also include covariate that capture the availability of infrastructure at the healthcare facility including hygiene levels (reference category is shortage of staff or equipment to maintain hygiene), storage availability (reference category is “No” or limited storage available), availability of electrical outlets (reference category is less than sufficient working electricals outlets), availability of distilled/sterile water (reference category is less than sufficient access to distilled/sterile water).

### Ethical Considerations

All respondents provided verbal informed consent to participate in the study. No personally identifiable information was used for any of the analysis. The assessments did not influence the time or place of staff responsible for care at facilities nor impose significant additional burden on patients or staff. The study protocol received approval from the Human Subjects Committee of the Institute of Financial Management and Research (IFMR IRB), Ref(PIRB00007107; FWA00014616; IORG0005894)

### Limitations of the Study

A retrospective assessment limits the ability to determine cause and effect which can attribute impact to a particular intervention. There was also limited flexibility in selection of the sample for the primary endpoint facilities survey with a focus on states that received greater support in terms of equipment allocation and distribution. This has implications on the generalizability of certain results identified under the intervention. There was insufficient information on total number of OCs and cylinders present at a facility to permit a baseline assessment. The survey tool focusses on the combined effect of OCs and cylinders. The two devices are likely to have varied long-term use cases.

## Results

A total of 450 healthcare facilities were surveyed, comprising of 22 (5%) COVID Care Centres, 157 Government Hospitals (Above the primary care level), 189 Government Primary Health Centres (PHC) / Community Health Centres (CHC) and 31 Private facilities. On average 26 oxygen devices (including OCs and Oxygen Cylinders) were delivered to Government Hospitals followed by 18.5 devices at COVID Care Centres (table 1). Private hospitals received 12.9 devices on average while PHC/CHC level Government facilities received 5.7 devices. In terms of location^5^, 315 (70%) of the facilities were located in rural or mostly rural districts while the remaining 135 (30%) of facilities were from urban or mostly urban districts. In terms of timeliness of receiving devices, 318 (71%) of devices were delivered during the peak^6^ of the second wave of COVID-19 in India. Over 133 (85%) of Government facilities received the devices during the peak while only 100 (53%) of PHC/CHC level facilities, located mostly in rural geographies received their devices during the peak months.

**Table 1:**
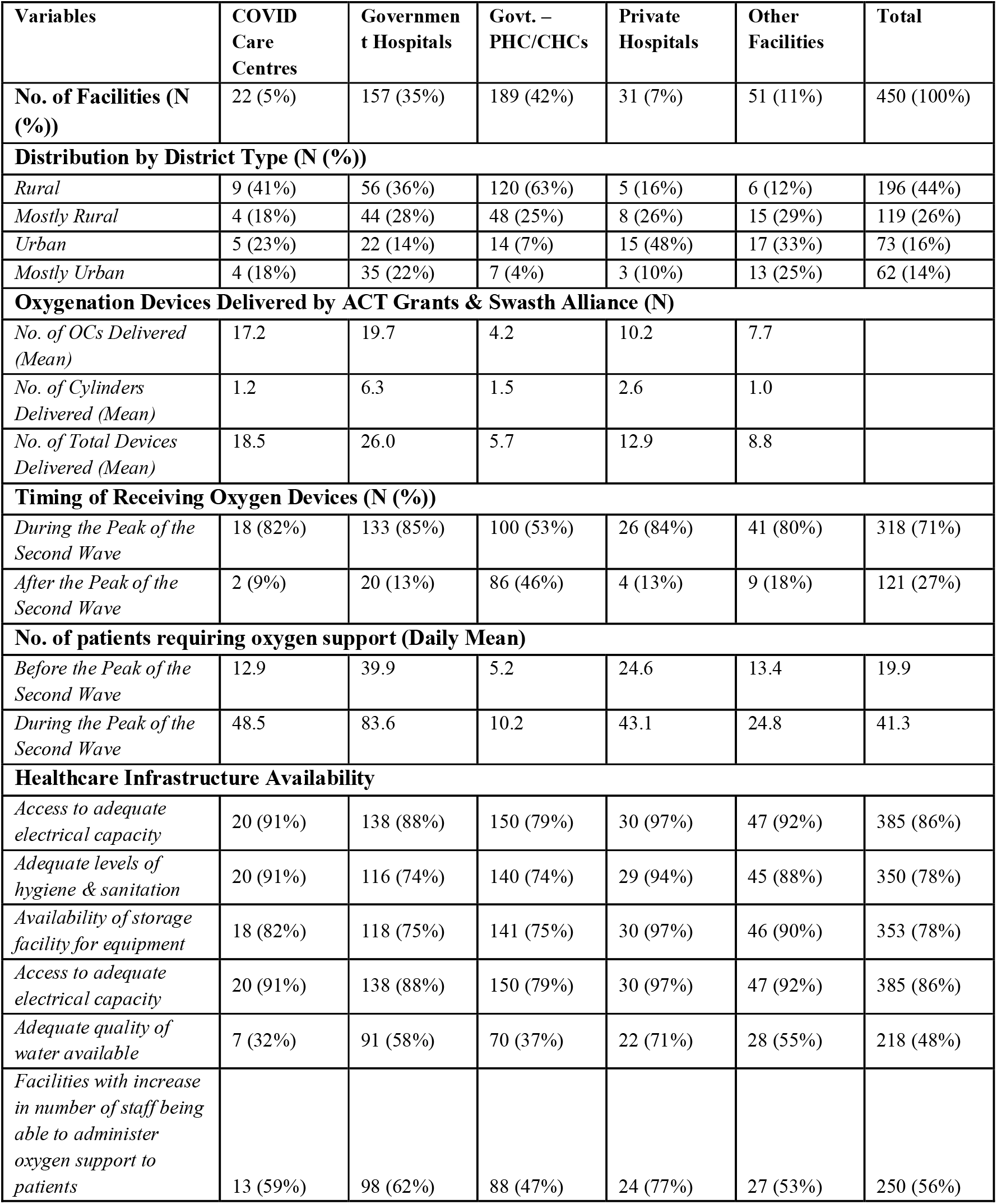
Healthcare Facility Characteristics (Sample)

| Variables | COVID Care Centres | Government Hospitals | Govt. – PHC/CHCs | Private Hospitals | Other Facilities | Total |
| --- | --- | --- | --- | --- | --- | --- |
| **No. of Facilities (N (%))** | 22 (5%) | 157 (35%) | 189 (42%) | 31 (7%) | 51 (11%) | 450 (100%) |
| **Distribution by District Type (N (%))** |  |  |  |  |  |  |
| *Rural* | 9 (41%) | 56 (36%) | 120 (63%) | 5 (16%) | 6 (12%) | 196 (44%) |
| *Mostly Rural* | 4 (18%) | 44 (28%) | 48 (25%) | 8 (26%) | 15 (29%) | 119 (26%) |
| *Urban* | 5 (23%) | 22 (14%) | 14 (7%) | 15 (48%) | 17 (33%) | 73 (16%) |
| *Mostly Urban* | 4 (18%) | 35 (22%) | 7 (4%) | 3 (10%) | 13 (25%) | 62 (14%) |
| **Oxygenation Devices Delivered by ACT Grants & Swasth Alliance (N)** |  |  |  |  |  |  |
| *No. of OCs Delivered (Mean)* | 17.2 | 19.7 | 4.2 | 10.2 | 7.7 |  |
| *No. of Cylinders Delivered (Mean)* | 1.2 | 6.3 | 1.5 | 2.6 | 1.0 |  |
| *No. of Total Devices Delivered (Mean)* | 18.5 | 26.0 | 5.7 | 12.9 | 8.8 |  |
| **Timing of Receiving Oxygen Devices (N (%))** |  |  |  |  |  |  |
| *During the Peak of the Second Wave* | 18 (82%) | 133 (85%) | 100 (53%) | 26 (84%) | 41 (80%) | 318 (71%) |
| *After the Peak of the Second Wave* | 2 (9%) | 20 (13%) | 86 (46%) | 4 (13%) | 9 (18%) | 121 (27%) |
| **No. of patients requiring oxygen support (Daily Mean)** |  |  |  |  |  |  |
| *Before the Peak of the Second Wave* | 12.9 | 39.9 | 5.2 | 24.6 | 13.4 | 19.9 |
| *During the Peak of the Second Wave* | 48.5 | 83.6 | 10.2 | 43.1 | 24.8 | 41.3 |
| **Healthcare Infrastructure Availability** |  |  |  |  |  |  |
| *Access to adequate electrical capacity* | 20 (91%) | 138 (88%) | 150 (79%) | 30 (97%) | 47 (92%) | 385 (86%) |
| *Adequate levels of hygiene & sanitation* | 20 (91%) | 116 (74%) | 140 (74%) | 29 (94%) | 45 (88%) | 350 (78%) |
| *Availability of storage facility for equipment* | 18 (82%) | 118 (75%) | 141 (75%) | 30 (97%) | 46 (90%) | 353 (78%) |
| *Access to adequate electrical capacity* | 20 (91%) | 138 (88%) | 150 (79%) | 30 (97%) | 47 (92%) | 385 (86%) |
| *Adequate quality of water available* | 7 (32%) | 91 (58%) | 70 (37%) | 22 (71%) | 28 (55%) | 218 (48%) |
| *Facilities with increase in number of staff being able to administer oxygen support to patients* | 13 (59%) | 98 (62%) | 88 (47%) | 24 (77%) | 27 (53%) | 250 (56%) |

On average, the daily number of patients requiring oxygen support was 19.9. This more than doubled to 41.3 patients daily during the peak months of the second wave in India. At Government facilities, the requirement for oxygen support nearly doubled from 39.9 patients before the second wave to 83.6 patients during the peak months of the second wave. Government hospitals reported being more affected by the increased demand of oxygen support compared to other types of healthcare facilities.

### Facility Infrastructure

Table 1 describes the infrastructure available at the surveyed healthcare facilities and the challenges to the long-term usage of oxygenation devices received by them. In terms of availability of adequate electrical infrastructure (in terms of functional electric outlets and uninterrupted power supply), 350 (86%) of facilities reported in the positive. Lack of distilled water for the oxygen concentrator was a bottleneck reported by over half 218 (48%) of facilities with COVID Care Centres and PHC/ CHC level facilities faring worse. 210 (17%) of facilities reported that there was an adequate availability of medical equipment repair technicians in case of any issues with the devices they received. While 24 (77%) of private facilities reported adequacy, only 70 (37%) of PHC/ CHC level facilities, located largely in rural or mostly rural districts had access to a sufficient number of technicians.

While private facilities 29 (94%) and COVID Care Centres 20 (91%) reported adequate levels of hygiene and sanitation, over one-fourth of Government facilities both at secondary and tertiary levels 116 (74%) and at the PHC/ CHC levels report not being able to maintain adequate levels of hygiene and sanitation at the facility. In a similar vein, while private facilities 30 (97%) and COVID Care Centres 15 (68%) reported adequate storage facilities for oxygenation equipment while over one of Government Hospitals lacked sufficient storage facilities with implications for long-term usage of these devices. Over 250 (56%) of facilities also reported that there has been an increase in the number of staff being able to administer oxygen support to patients since the beginning of the second wave.

### Utility of Oxygen Equipment in alleviating supply shortages in the short-run and long-run

#### Effect of additional delivery of OCs and cylinders on oxygen demand in the short run

A majority of (54%) facilities reported that distribution of oxygenation devices helped meet their oxygen demand (Table 2). 67% of the surveyed private hospitals and 64% of covid care centers reported to have benefitted from receiving these oxygenation devices in the short run in meeting the oxygen demand. The bivariate analysis associations showed that the timely availability of technicians, average load of COVID-19 patients during the second wave and timely availability of oxygenation devices such as OCs were predictors of additional oxygenation devices having a significant impact on meeting the oxygen demand at the facility (Figure 1). The multivariate analysis confirmed that facilities with adequate availability of medical equipment repair technicians (AOR: 1.77, CI: 1.02-3.08, p-value: 0.04) and those receiving oxygenation devices before the start of the second wave (AOR: 2.60, CI: 1.04-6.49, p-value: 0.04) were at higher odds of meeting the oxygen demand at the facility with the delivery of additional oxygenation devices.

**Table 2:** Utility of Devices in alleviating oxygen supply shortages

| Variables | COVID Care Centres | Government Hospitals | Government – PHC/CH Cs | Private Hospitals | Other Facilities | Total |
| --- | --- | --- | --- | --- | --- | --- |
| **Number of facilities** | 22 (5%) | 157 (35%) | 189 (42%) | 31 (7%) | 51 (11%) | 450 (100%) |
| **Increased capacity to admit non-COVID patients with oxygen needs in future (N (%))** |  |  |  |  |  |  |
| *After the Second Wave (Long-Run)* | 9 (41%) | 91 (58%) | 107 (57%) | 14 (45%) | 18 (35%) | 239 (53%) |
| **Reduced administrative load on staff to organize oxygen support (N (%))** |  |  |  |  |  |  |
| *After the Second Wave (Long-Run)* | 6 (27%) | 35 (22%) | 45 (24%) | 13 (42%) | 9 (18%) | 108 (24%) |
| **Effect of additional delivery of devices on oxygen demand (short-run) (N (%))** |  |  |  |  |  |  |
| *Significantly Decreased or fully met oxygen demand* | 14 (64%) | 85 (55%) | 96 (50%) | 21 (67%) | 26 (51%) | 242 (54%) |

**Figure 1:**
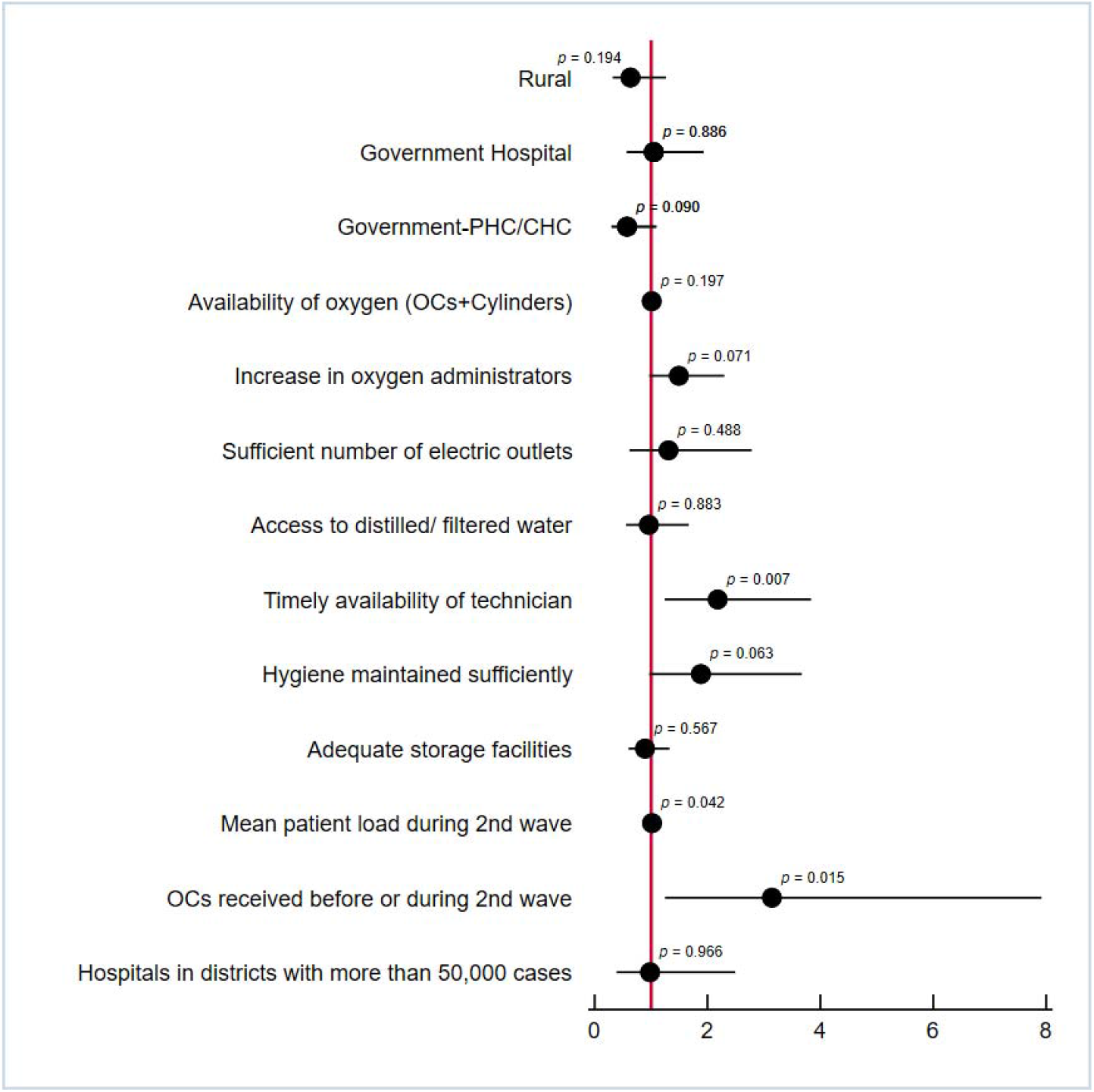
Bivariate analysis for additional OCs and cylinders on oxygen demand in the short-run

#### Expected increase in capacity of the facility in the long-run

Around 239 (53%) facilities expected an increase in the capacity of the facility to admit patients with non-COVID illnesses (that require oxygen support) in the long run. Perceived capacity to admit patients with non-COVID-19 illnesses, in the long run, was highest in government hospitals (58%) and government PHCs/CHCs (57%), compared to other facilities (Table 2). As per the bivariate (Figure 2) and the multivariate analyses, the potential predictors of the expected increase in the capacity of the facility to admit non-COVID-19 patients that required oxygen in the long run included type of facility such as government hospitals (AOR:1.95, CI: 1.22 – 3.11, p-value: 0.005) and government PHC/CHC (AOR: 2.58, CI: 1.44-4.63, p-value: 0.001), availability of oxygenation devices (AOR: 1.01, CI: 1.00-1.02, p-value:0.015) and sufficient number of electric outlets (AOR: 2.64, CI: 1.46-4.80, p-value: 0.001).

**Figure 2:**
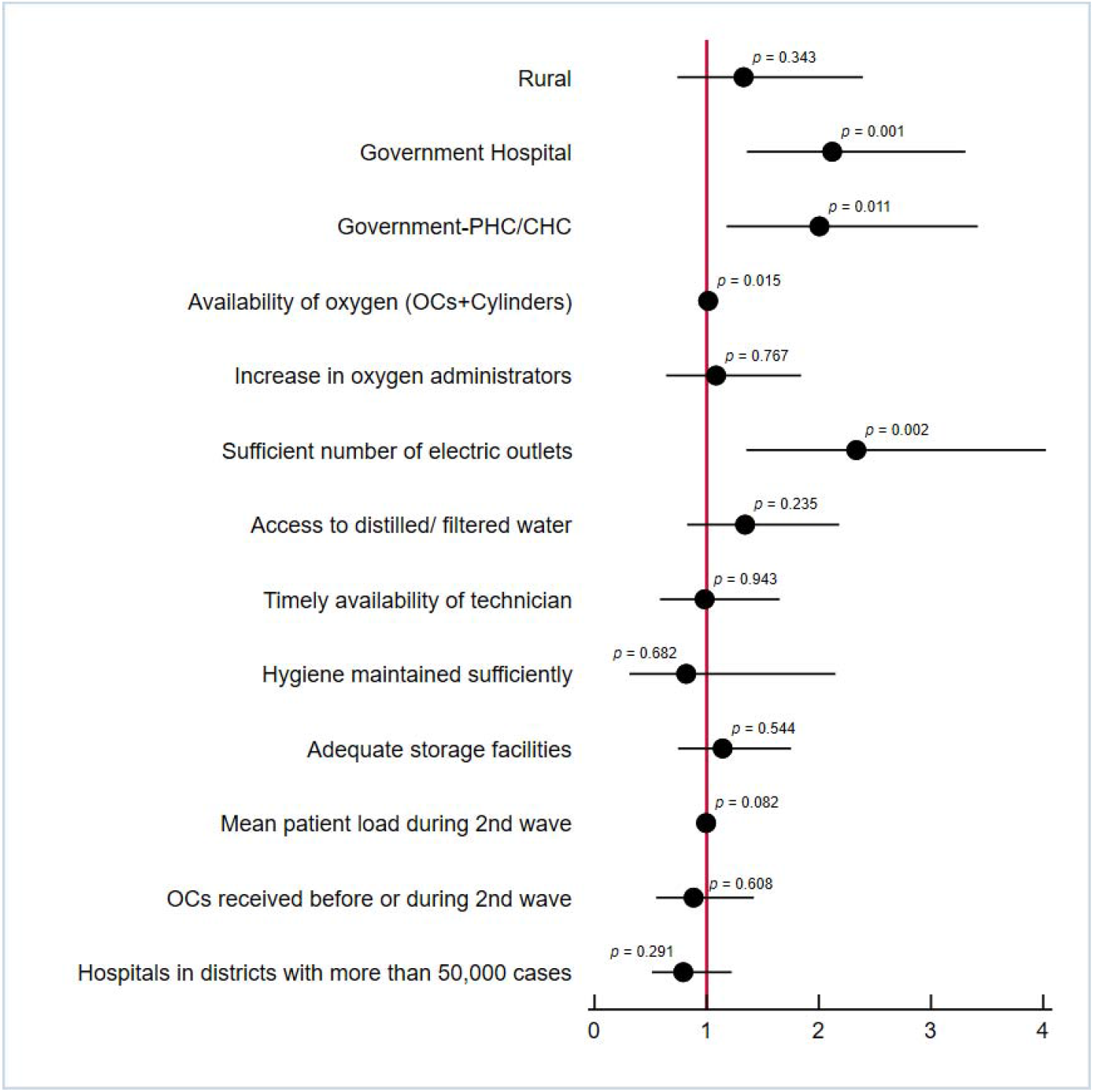
Bivariate analysis for additional OCs and cylinders on expected increase in capacity of the facility in the long-run

#### Expected reduced administrative load on staff to organize oxygen support in the long-run

In terms of reducing the administrative work for staff to organize oxygen support, 42% of the surveyed private hospitals perceive a higher impact of oxygenation devices in the long-run, compared to 24% of the overall surveyed facilities (Table 2). The bivariate and multivariate analyses showed that facilities with sufficient number of electric outlets (AOR:2.44, CI: 1.34-4.45, p-value: 0.003) and facilities that increased the number of staff members that were able to administer oxygenation devices in the beginning of the second wave (AOR: 2.02, CI: 1.21-3.37, p-value: 0.007) were at higher odds of expecting a reduction in the administrative load on their staff to organize oxygen support in the long run (Figure 3).

**Figure 3:**
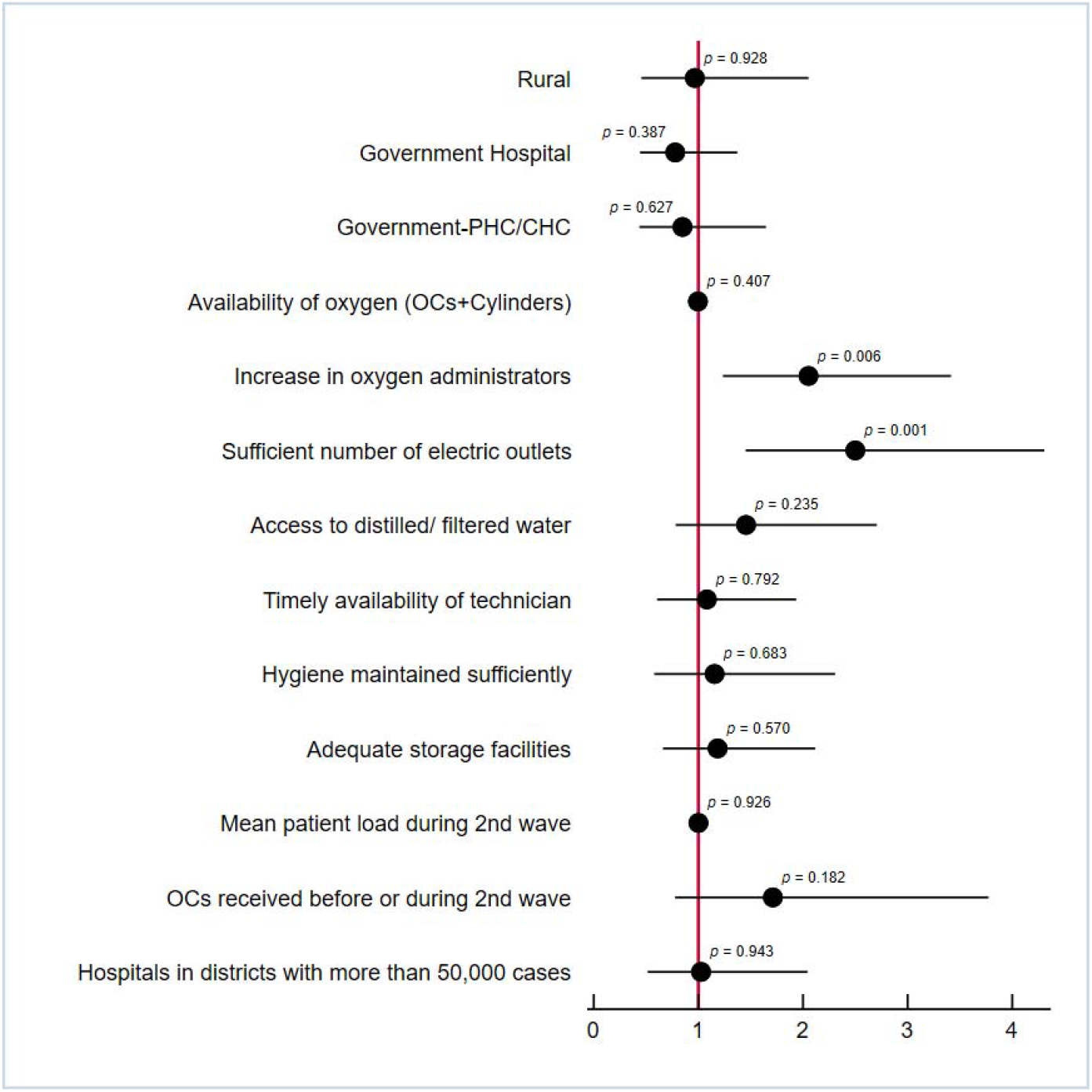
Bivariate analysis for additional OCs and cylinders on expected reduced administrative load on staff to organize oxygen support in the long-run

## Discussion

Recent literature assessing efforts globally in addressing the acute oxygen crisis triggered by multiple waves of COVID-19 have highlighted the high level of complexity associated with managing such shortfalls. Ensuring increased availability of oxygen at the level of healthcare facilities is not sufficient in enabling sustainable and efficient usage. A majority of (54%) facilities reported that distribution of oxygenation devices helped meet their oxygen demand during the peak months of the second wave of COVID-19 in India. We find that the ability to meet the oxygen demand at the facility is associated with facility level factors, availability of trained manpower as well as infrastructural sufficiency.

Timeliness in receiving oxygen devices before or during the onset of the surge in demand enabled facilities to better allocate and efficient use the available oxygen for COVID patient caseloads. Availability of oxygen also shows increase in the ability to treat non-COVID related patients especially in Government facilities and PHC/ CHC level facilities with low-resource constraints. Making oxygen devices available in government facilities is crucial in developing an inclusive healthcare system as the poorest quintile of the population has a higher dependency on public healthcare facilities for in-patient support compared to other income classes^7^.

Investing in training personnel to administer and maintain oxygen can provide efficiency gains in the longer term and reduce administrative burden. Our findings suggest that facilities that were able to increase the number of staff able to administer oxygen devices were more likely to be able to meet their oxygen needs. Long-term maintenance of devices with timely availability of biomedical repair technicians is an important predictor of facilities being able to meet their oxygen demand. Biomedical engineer training has been slow to develop to help address this. However, some Government programmes have begun to recognise the complex nature of biomedical technology and associated healthcare equipment management.

While investing in personnel may take years to bear fruition, government agencies and private entities donating or subsidising these devices must carefully consider long-term maintenance budgets when assessing the feasibility and impact of their contributions, particularly regarding the ongoing cost burden of related infrastructural facilities. These include funds to ensure maintenance of adequate electrical load and outlets which are important for the operation of oxygen concentrators in particular. Additionally, there are several cultural and bureaucratic factors particularly in the public healthcare domain that can significantly affect maintenance of equipment. Under-reporting breakdown of medical devices is common, in the absence of maintenance funds and repair technicians. Creating an enabling environment may help to improve device lifetime and associated cost burdens.

Another aspect is the sourcing of oxygen equipment, which remains problematic and requires further study. Medical equipment is largely produced in developed, high income country contexts, even those specifically designed for low-resource settings and environment condition in the country importing these devices (Marks, Thomas, & Bakhet, 2019). For instance, non-availability of operating manuals in local vernacular can be an impediment in properly using and maintaining this equipment at healthcare facilities. The facility-specific nature of these challenges however, limits the generalizability of specific solutions.

## Conclusion

We carried out a survey in health facilities which received oxygenation devices during the second wave of COVID-19 in India on the reported short-run and expected long-run utility of these devices. Our findings suggest that these devices helped meet the short run oxygen demand and are likely to increase the hospital capacity in the long-run to admit non-COVID patients. However, these devices can be fully utilized to meet both short and long run oxygen shortages only if there are skilled technicians and oxygen administrators to help in use and repair of these devices. It will also require creation of supportive infrastructure such as sufficient number of electrical outlets in health facilities, which is a particularly relevant result for the resource constrained LMIC setting.

## Data Availability

All data produced in the present study are available upon reasonable request to the authors

## Contributors

DB developed the data analysis plan and led the data and writing. VK developed the survey instrument tool and managed the data collection, management and quality check. SB, AN and NJ had full access to all the data in the study and had final responsibility for the decision to review and submit the manuscript for publication. DB and VK wrote the first draft of the manuscript. All authors contributed to the final version of the paper.

## Data sharing statement

De-identified and anonymized healthcare facility-level primary data used for the analysis can be made available on request.

## Declaration of Conflict of Interests

We declare no competing interests.

## Acknowledgements

We would like to thank Dr. Ramanan Laxminarayan for his comments and suggestions which improved the quality of the paper. We would like to thank Abhik Banerji and Srishti Goel from One Health Trust, India for helping with the data analysis and the field operations team at LEAD at Krea University for conducting the primary data collection and data management. We would like to thank the study participants for giving off their time to complete the primary survey. We sincerely thank the reviewers for their comments and suggestions which improved the quality of the paper.

## Footnotes

1 Source: Directorate General of Commercial Intelligence and Statistics (DGCIS)

2 https://www.covid19india.org/

3 https://mppn.org/paises_participantes/india/

4 Maintained internally by ACT Grants with information of the distribution of oxygen devices at a facility level.

5 *Rural: More than 75% rural population* *Majority Rural: 50-75% of rural population* *Majority Urban: 50-75% urban population* *Urban: More than 75% urban population*

6 To define the peak, we have plotted the daily active caseload data for the entire second wave period and then identified the peak based on observations that correspond to the 75th percentile or higher in the caseload distribution (corresponds roughly to about a 35–40-day period for most districts). This gives us bounds for the peak.

7 Source: NSS Health Survey 2017-18

## Notes

### Competing Interest Statement

The authors have declared no competing interest.

### Funding Statement

This study did not receive any funding.

### Author Declarations

The study protocol received approval from the Human Subjects Committee of the Institute of Financial Management and Research (IFMR IRB), Ref(PIRB00007107; FWA00014616; IORG0005894)

## References

Bakare, A. (2020). Providing oxygen to children and newborns: a multi-faceted technical and clinical assessment of oxygen access and oxygen use in secondary-level hospitals in southwest Nigeria. International Health.

Baker, T. S. (2020). Essential care of critical illness must not be forgotten in the COVID-19 pandemic. The Lancet, 395:1253–4.

Bonnet, L., Carle, A., & Muret, J. (2021). In the light of COVID-19 oxygen crisis, why should we optimise our oxygen use? 2021. Anaesthesia Critical Care & Pain Medicine.

Bradley, B., Light, J., Ebonyi, A., N’Jai, P., Ideh, R., Ebruke, B., … Howie, S. (2016). Implementation and 8-year follow-up of an uninterrupted oxygen supply system in a hospital in the Gambia. Int Journal of Tuberculosis and Lung Disease.

Dauncey JW, O.-O. P. (2019). Healthcare-provider perceptions of barriers to oxygen therapy for paediatric patients in three government-funded eastern Ugandan hospitals; a qualitative study.. BMC Health Services Research.

Duke, T., Graham, S., & Cherian, M. (2010). Oxygen is an essential medicine: a call for international action. International Journal of Tuberculosis and Lung Disease, 14:1362–8.

Duke, T., Peel, D., Wandi, F., Subhi, R., Sa’avu, M., & Matai, S. (2010). Oxygen supplies for hospitals in Papua New Guinea: a comparison of the feasibility and cost-effectiveness of methods for different settings. Papua New Guinea Medical Journal, 53: 126–138.

Goh. (2018). 2018. A government-led approach to increasing access to oxygen in Nigeria. Retrieved from CHAI: https://www.clintonhealthaccess.org/government-led-approach-accessing-oxygen-nigeria/

Graham HR, M. J. (2021). Graham HR, Maher J, Bakare AA, Nguyen CD, Ayede AI, Oyewole OB, Gray A, Izadnegahdar R, Duke T, Falade AG. Oxygen systems and quality of care for children with pneumonia, malaria and diarrhoea: Analysis of a stepped-wedge trial in Nigeria. PLoS One.

Guardian, T. M. (2021, May). After India: the countries on the brink of another Covid oxygen crisis.. Retrieved from The Mail & Guardian: https://mg.co.za/health/2021-05-25-after-india-the-countries-on-the-brink-of-another-covid-oxygen-crisis/

Howie, S. (2010). Meeting oxygen needs in Africa: An options analysis from the Gambia. Bulletin of the World Health Organization.

IVAC, I. V. (2020). Pneumonia & diarrhea progress Report. International Vaccine Access CenterJohns Hopkins Bloomberg School of Public Health.

Macnamara, K. (2020). ‘Suffering, gasping’: experts warn of oxygen shortages in poorer virus-threatened nations. Retrieved from Barrons: https://www.barrons.com/news/suffering-gasping-experts-warn-of-oxygen-shortages-in-poorer-virus-threatened-nations-01587527706

Marks, I., Thomas, H., & Bakhet, M. (2019). Medical equipment donation in low-resource settings: a review of the literature and guidelines for surgery and anaesthesia in low-income and middleincome countries. BMJ Global Health.

Meara, J. e. (2015). Global surgery 2030: evidence and solutions for achieving health, welfare, and economic development. The Lancet, 386: 569–624.

Stein, F., Perry, M., & Banda, G. (2020). Oxygen provision to fight COVID-19 in sub-Saharan Africa. BMJ Global Health.

Thadani, A. (2021). Preventing a Repeat of the COVID-19 Second-Wave Oxygen Crisis in India. ORF.

WHO. (2021). COVID-19 clinical management: living guidance, 25 January 2021. World Health Organization.

Zhu. (2020). Coronavirus exposes Africa’s oxygen problem. Retrieved from New Humanitarian: https://www.thenewhumanitarian.org/news/2020/04/16/Africa-oxygen-problem-coronavirus

